# Effectiveness of a COVID-19 contact tracing app in a simulation model with indirect and informal contact tracing

**DOI:** 10.1101/2023.06.15.23291010

**Authors:** Ka Yin Leung, Esther Metting, Wolfgang Ebbers, Irene Veldhuijzen, Stijn P. Andeweg, Guus Luijben, Marijn de Bruin, Jacco Wallinga, Don Klinkenberg

## Abstract

During the COVID-19 pandemic, contact tracing was used to identify individuals who had been in contact with a confirmed case so that these contacted individuals could be tested and quarantined to prevent further spread of the SARS-CoV-2 virus. Many countries developed mobile apps to find these contacted individuals faster. We evaluate the epidemiological effectiveness of the Dutch app CoronaMelder, where we measure effectiveness as the reduction of the reproduction number R. To this end, we use a simulation model of SARS-CoV-2 spread and contact tracing, informed by data collected during the study period (December 2020 - March 2021) in the Netherlands. We show that the tracing app caused a clear but small reduction of the reproduction number, and the magnitude of the effect was found to be robust in sensitivity analyses. The app could have been more effective if more people had used it, and if time intervals between symptom onset and reporting of contacts would have been shorter. The model used is novel as it accounts for the clustered nature of social networks and as it accounts for cases informally alerting their contacts directly after symptom onset, without involvement of health authorities or a tracing app.

## 1. Introduction

In most European countries, the main non-pharmaceutical control measures against the spread of SARS-CoV-2 in the first two years were aimed at reducing contacts or risk of transmission per contact on population level, for instance by closing schools or restaurants, working from home, keeping distance, or wearing face masks. Some of these measures may have had a negative impact on the society, in particular to mental health and the economy (Ashraf, 2020; Van den Boom, 2022). For that reason, control measures focusing on isolating only infected individuals and quarantining their contacts were preferable.

In infectious disease control, contact tracing is a widely applied strategy to find potentially infectious individuals, quarantine and test them. Traditionally, contact tracing is done manually by public health professionals and relies on the ability and willingness of an individual to recall and report close contact events (to be referred to as contacts), and of the contacted persons (to be referred to as contactees) to follow the quarantine and testing rules. Modelling studies show that the effectiveness of contact tracing strongly depends on characteristics of the infection such as the incubation period and generation time, and can be very sensitive to delays in the tracing process such as waiting for test appointments or the time it takes for health authorities to call cases, ask for contacts, and then inform contactees, which increase the time between transmission and starting quarantine (Fraser et al., 2004; Hellewell et al., 2020; Klinkenberg et al., 2006; Kretzschmar et al., 2021; Muller et al., 2000).

To improve the process of contact tracing, various countries developed contact tracing apps which informed users that they were contacted by someone who was tested positive for SARS-CoV-2.

These apps were meant to reduce the time interval between symptom onset of a case and the identification of their contacts with shorter time delays, and to trace contactees not known to or recalled by the case. Early modelling studies (Cencetti et al., 2021; Ferrari et al., 2021; Kucharski et al., 2020; Kurita et al., 2021) suggested a potentially relevant contribution of such apps to COVID-19 control. Post-implementation analyses in the UK showed that many infections were indeed prevented by their contact tracing app (Ferretti et al., 2020). However, also with the digital contact tracing app, control by social distancing measures remained necessary.

Since the start of the pandemic many studies have been conducted on contact tracing with contact tracing apps (Braithwaite et al., 2020; Jenniskens et al., 2021). Mathematical modelling studies have applied several types of models, ranging from compartmental models (Ge et al., 2021; Nuzzo et al., 2020) and branching process models (Bradshaw et al., 2021; Kretzschmar et al., 2020), to agent-based models (Cencetti et al., 2021; Kucharski et al., 2020; Quilty et al., 2021; Scott et al., 2021). A consistent conclusion across studies is that contact tracing apps can contribute to the control of an epidemic, but the extent of the impact is very sensitive to the percentage of the population using the app and the delays in the process of tracing and isolating infectious contacts. In some studies (e.g. (Kretzschmar et al., 2020)) the importance of short delays of steps in the contact tracing process is emphasized whereas in other studies (e.g. (Kucharski et al., 2020)) the emphasis is on the importance of having a high proportion of cases to isolate and a high proportion of their contacts to be successfully traced.

In 2021, the National Institute for Public Health and the Environment (RIVM) was asked for a model-based evaluation of the epidemiological effectiveness of the Dutch contact tracing app CoronaMelder. Here we present the model and the evaluation. We measure effectiveness of contact tracing as the resulting reduction in the reproduction number R, which is defined as the mean number of secondary cases per primary case (see (Klinkenberg, 2021)). The model is informed with data from surveillance sources and behavioural surveys during the first months of 2021 in the Netherlands. The model itself includes two features that are novel. First, we account for the clustered nature of social contact networks, which is relevant as it increases the probability that infected individuals can be notified by a case in their social network without having been infected by that case. Second, we explicitly account for notification of contacts by cases themselves, without involvement of health authorities or a tracing app. We estimate the additional effect of manual contact tracing over this informal notification, and the additional effect of the contact tracing app over the combined effect of informal notification and manual contact tracing.

## 2. Methods

### 2.1 Data sources

Model parameters were estimated from data on the Dutch COVID-19 epidemic as much as possible. Epidemiological data from three surveillance systems were used:

- Osiris: the national case notification registry containing all confirmed COVID-19 cases (Ward et al., 2005)
- CoronIT: tests carried out by the Municipal Health Services, from all public testing facilities
- HPZone: all contact tracing data

Behavioural data from three large studies were used:

- CGU: questionnaires about beliefs and adherence to measures, ran by the Corona Behavioural Unit of the RIVM (RIVM, 2021)
- LISS: questionnaires about the tracing app, beliefs and adherence to measures, with an existing longitudinal Social Sciences cohort panel (Van der Laan, 2021)
- PanelClix: questionnaire about the tracing app and how it is used, with an existing internet panel (Ebbers, 2021)

### 2.2 Epidemiological situation

The evaluation was done for the period between 1 December 2020 and 31 March 2021, when vaccination was not yet available, and lockdown restrictions were in place such as keeping distance, working from home, and closed businesses. During that period, all testing was done by public testing facilities (no self-administered antigen tests). People were allowed to get tested when they had symptoms, or five days after contact with a positive case identified by manual contact tracing. Notification of close contacts through the contact tracing app was done through the health authorities: when cases were approached by the health authorities for manual contact tracing, they were asked whether they used the app, and if so, they were provided a key by which they could activate the app to notify their contactees. Notified individuals were advised by the app to quarantine.

### 2.3. Model formulation and analysis

The effectiveness of contact tracing was expressed in terms of a percentual reduction of the reproduction number R. This reproduction number was determined with a simulation model for SARS-CoV-2 transmission and contact tracing. This was done for several parameter sets: the baseline parameter set, parameter sets for sensitivity analyses, and parameter sets for scenarios to explore improvements to the app and its use.

For each parameter set we simulated 10,000 transmission trees (outbreaks starting with one index) of at most 10 generations which is equivalent to about 1.5 months of virus spread starting with an incidence rate of 2,500 cases during four days (with a mean generation interval of 4 days). In the simulations all cases and transmission links are known, therefore it is possible to calculate the mean number of secondary cases across all transmission chains, which is the reproduction number R. The epidemiological effectiveness of testing, case isolation, contact tracing and quarantine was determined by applying sets of control measures to the 10,000 simulated transmission trees, removing cases if they would have been prevented, and recalculating the reproduction number.

The control measures were applied at different levels:

1. no control
2. testing and informal tracing: cases test and go into quarantine because of symptoms; cases go into isolation because of a positive test result; contactees test and go into quarantine because of informal contact tracing
3. manual tracing: contactees test and go into quarantine because of manual contact tracing (in addition to 1)
4. tracing app: contactees test and go into quarantine because of an app notification (in addition to 1 and 2)

The total effectiveness for a parameter set was determined as the reduction in *R* from no control to control level 3. The contributions of informal tracing, manual tracing, and app-based tracing separately were determined as the reductions in *R* from no control to level 1, from level 1 to level 2, and from level 2 to level 3, respectively.

Specific details of the Dutch testing policy were modelled explicitly: in control level 1, testing of contacts was only done when they were symptomatic and in control levels 2 and 3, testing of contacts was done when symptomatic or five days after infection (whichever was earlier). Of course, in all cases testing, isolation and quarantine were also conditional on adherence. See also section 2.3.2.

The simulation model consisted of three layers, explained in more detail below. The first layer was the transmission model, used to create transmission trees without contact tracing interventions. This resulted in a chronological description for all cases of who had contacted whom, and when symptoms had started (if any). The second layer was the behaviour model, used to assign behavioural parameters to each case that determine what actions they are willing to take to help control the epidemic. Actions are testing or isolation/quarantine, and are only taken when triggered by symptom onset, a positive test result, or notification through contact tracing. The behaviour model is also used to determine who is willing to use the contact tracing app (app use). The third layer was the logistics model, used to apply testing and tracing to the transmission trees with all relevant delays in the test-and-trace process, and the probabilities by which contacts are traced. The second- and third layer were tailored to the Dutch situation in the study period, e.g. testing rules and notification processes. Further details are provided in the model description below.

#### 2.3.1 Transmission: epidemic outbreak and social network with clustering

Transmission trees were simulated with a branching process with negative binomial offspring distribution, up to a maximum number of 10 generations after the index case. The offspring distribution is defined by a mean *Rref* (the reference reproduction number, i.e. without testing and tracing) and a shape *k*, which determines the variation and therefore the possibility of superspreading. The reproduction number *Rref* in the simulation is without isolation or quarantine, but should be interpreted in the presence of population-level control measures such as keeping physical distance, working from home, and closed businesses.

For each case a time of infection was sampled, as well as a time of symptom onset, with a certain fraction of cases without symptoms (see also Table 1). Starting with infection time 0 for the index case of the branching process, the infection times for the subsequent cases were always one random generation interval later. The times of symptom onset were equal to the infection times plus a randomly sampled incubation period. Generation interval and incubation period were modelled with gamma distributions.

**Table 1:** Parameters of the transmission model

| Description | Baseline value <sup>a</sup> | Source <sup>b</sup> |
| --- | --- | --- |
| Reproduction number $R_{ref}$ | 1.3 | <sup>c</sup> |
| Contacts |  |  |
| Superspreading coefficient $k$ (shape parameter of the negative binomial distribution with mean $R_{ref}$) | 0.1 | (Kirkegaard and Sneppen, 2021; Susswein, 2020) |
| Clustering probability $c$ | 0.2 | (Eames and Keeling, 2003; Kissler, 2020; Kucharski et al., 2018) |
| Course of an individual infection |  |  |
| Percentage of infections with symptoms | 70% | (Buitrago-Garcia et al., 2020; McDonald et al., 2021) |
| Mean incubation period | 5 days | (Backer et al., 2020; Cheng, 2020) |
| Mean generation interval | 4 days | Osiris |

<sup>a</sup> Value of the baseline parameter set; see Supplement S2 for values used in sensitivity analyses and future scenarios
<sup>b</sup> Literature reference or dataset: Osiris = case notification registry; see Supplement for more detail
<sup>c</sup> The exact value of $R$ is not essential to calculate a relative reduction. A value larger than one is necessary for an outbreak to enable computations. A value close to one is most representative for the evaluation period.

We accounted for the local clustering of social contact networks by adding edges to the transmission trees that represent contacts that had not led to transmission but which may be used for contact tracing. These edges were placed such that networks with triangles were formed, in three steps:

- each pair of individuals that had a contactee in common was linked with probability c.
- each pair of individuals that had a contactee in common because of step 1, was linked with probability *c*^2^.
- Each pair of individuals that had a contactee in common because of step 2, was linked with probability *c*^3^.

Parameter values with justification are listed in Table 1, with more detail in Supplement S1.

#### 2.3.2 Behaviour: adherence to control measures

Behaviour was parameterised by a set of probabilities of adherence to testing, isolation, quarantine and app use (the behaviours), in response to symptoms, a positive test result, any type of contact tracing, or availability of a contact tracing app (the triggers). All individuals in the simulated outbreaks were randomly assigned three adherence levels between 0 and 1: one level related to testing, one related to isolation and quarantine, and one related to app use. These levels determined the behaviour of an individual in response to a trigger, by comparing their adherence level to a probability. For instance, if app use was 30%, the 30% individuals that were assigned the highest app-use adherence level (app-use level > 0.7) used the contact tracing app. By using adherence levels instead of random behaviours for each trigger-behaviour combination, we created consistency in each individual’s behaviour across triggers and parameter sets. The probabilities for each trigger- behaviour combination are listed in Table 2, with more detail in Supplement S1.

**Table 2.** Parameters of the behaviour model. Adherence levels of actions in response to triggers

| Description | Baseline value <sup>a</sup> | Source <sup>b</sup> |
| --- | --- | --- |
| Scheduling a test in response to... |  |  |
| ...symptoms (own initiative) | 50% | CGU |
| ...informal tracing notification | 68% | CGU |
| ...manual tracing notification | 90% | CGU |
| ...tracing app notification | 81% | CGU, LISS, PanelClix |
| Isolation in response to a positive test | 90% | CGU |
| Quarantine in anticipation of test outcome, in response to... |  |  |
| ...symptoms (own initiative) | 50% | CGU |
| ...informal tracing, manual tracing or tracing app (with symptoms) | 75% | CGU |
| ...informal tracing, manual tracing or tracing app (without symptoms) | 50% | CGU, PanelClix |

<sup>a</sup> Value of the baseline parameter set; see Supplement S2 for values used in sensitivity analyses and future scenarios
<sup>b</sup> Dataset: CGU = questionnaires by Corona Behavioural Unit; LISS = questionnaires in LISS Social Sciences cohort panel; PanelClix = questionnaire in PanelClix internet panel; see Supplement for more detail

#### 2.3.3 Logistics: delays and tracing efficiency

The process of testing and informing contacts, potentially followed by quarantine and isolation, is described by a cycle of two steps, initiated by a trigger step. Most actions are conditional on adherence, as described above.

1. The trigger step is the period between symptom onset and scheduling a test. During this step, there is no quarantine.
2. The first step of the cycle is the period between scheduling the test and receiving the test result. During this period, the actual test is taken (not explicitly modelled). This first step may be the start of the case’s quarantine. Upon receiving the positive test result, the case may become an index in the tracing process.
3. The second step of the cycle is the period between receiving the positive test result of the index and the scheduling of tests by his/her contacts. This second step may be the start of isolation of the index. During this period, the contacts may be informed by the modes of contact tracing that are in place, with probabilities depending on the effectiveness of recalling and notifying by the index, the contact tracing app, and the authorities.

The parameter values of the logistics model are listed in Table 3, with more detail in Supplement S1.

**Table 3.**
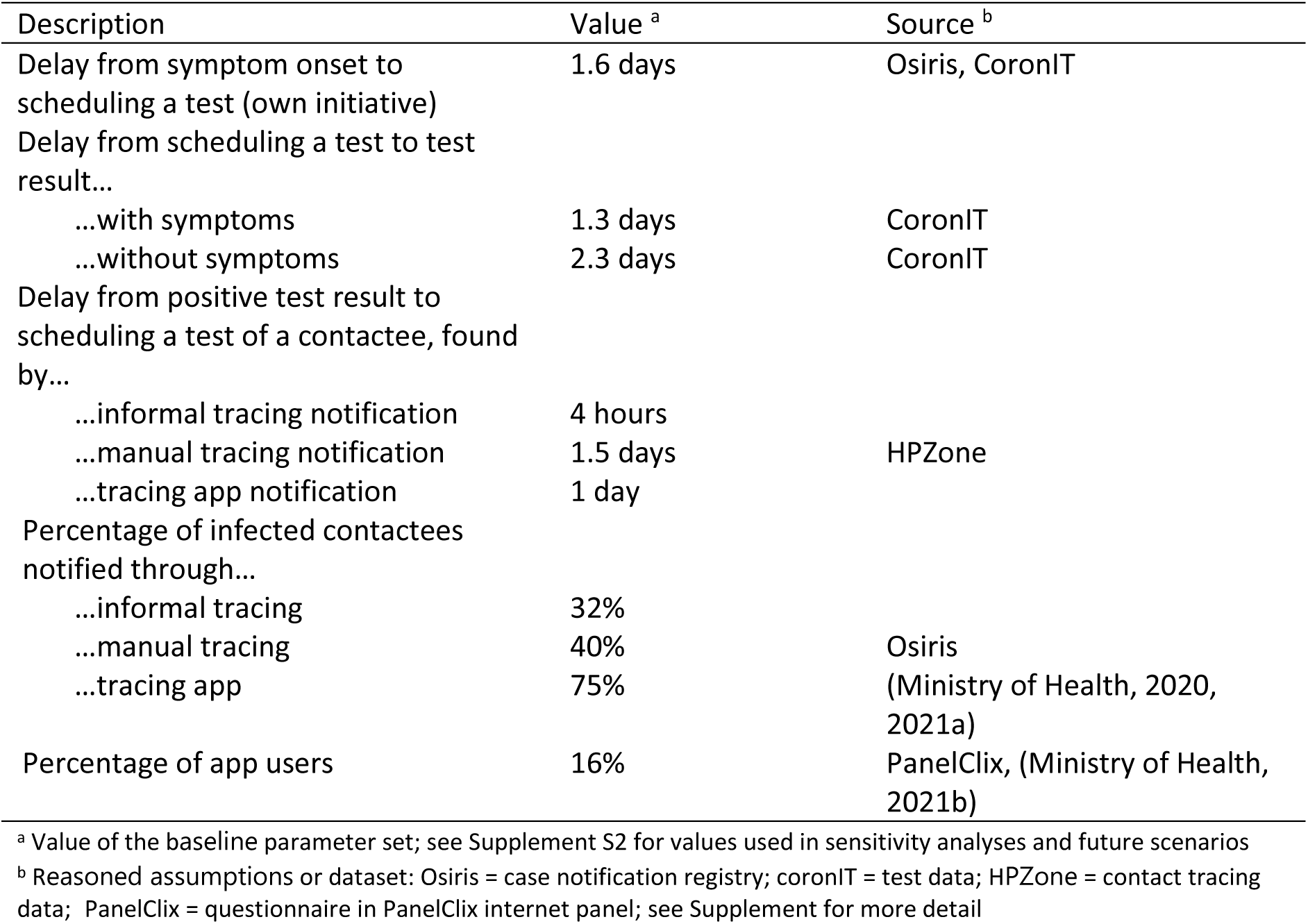
Parameters of the logistics model

#### 2.3.4 Applying testing and tracing to the simulated outbreaks

Implementation of control was simulated by first starting the test-and-trace cycle once for each case *i*, resulting in quarantine and isolation times *t_ij_* for all cases *j* (*t_ij_* =oo if *j* is never reached from *i*). From these times, min*_i_* (*t_ij_*) was determined as the end of the contact period for case *j* (i.e. minimum over all cases i), and all secondary cases of case *j* after these end times were removed, plus all cases from the generations thereafter. From all resulting transmission trees, the numbers of cases *C_g_* in each generation *g* were added, and the geometric average reproduction ratio was calculated over 7 generations as *R* (*C*_9_ *C*_2_)^1/7^ we skipped the initial and final generation to reduce edge effects.

In sensitivity analyses and scenarios in which only parameters concerning behaviour or logistics were changed (Tables 2-3), the same set of 10,000 transmission trees and individual adherence levels were used as with the baseline parameter set. Only when parameters for the transmission tree model were changed (Table 1), new transmission trees were simulated.

The code implementation of the model described in this paper was done in R version 3.6.0 (2019-04- 26) (Team, 2019), platform: x86_64-redhat-linux-gnu (64-bit). All code for the model and the analysis is published on https://github.com/rivm-syso/cm-evaluation.

#### 2.3.5 Baseline parameter set, sensitivity analyses, and future scenarios

The baseline parameter set (Tables 1-3) was estimated from data or obtained from literature, and was meant to best represent the situation in the Netherlands between December 2020 and March 2021, as described in Section 2.2.

In the baseline parameter set, 10% of the population would never test and therefore never go into isolation or quarantine, and 16% would use the app. In the baseline simulations, these behaviours were randomly drawn for each individual, but behaviours may be clustered in social networks. We explored the effect of clustering by dividing the population into two equal parts, one with higher adherence (5% never testing, 24% app use) and one with lower adherence (15% never testing, 8% app use), and determining the effectiveness in the two subpopulations.

Sensitivity analyses were carried out with alternative parameter sets or assumptions, exploring the main uncertainties of the baseline parameters (details in Supplement S2): a lower reproduction number, a longer generation interval, less or more superspreading, less or more clustering in the tracing network, more asymptomatic infections, lower tracing probabilities, and a smaller or larger proportion of the population not adhering to anything. In addition, we considered the possibility that adherence was socially clustered, so that a mean adherence level actually results high adherence in some transmission trees, and low adherence in other transmission trees. For this, we calculated the effectiveness in low-adherence and high-adherence transmission trees.

Future scenarios were evaluated to explore the potential contribution of the contact tracing app. First, we considered a change in policy in letting app users notify their contacts directly in the app and not through the health authorities. Second, we explored the effect of the likely reduced effectiveness of manual tracing if the lockdowns (in place during the evaluation period) would be lifted. This was expected to increase contact rates, especially with strangers, making informal and manual contact tracing less effective. In another future scenario, it was even considered to completely terminate formal manual contact tracing. Finally we considered the possibility of increasing the percentage of the population using the app (by campaigns). All these developments were evaluated through simulations of several parameter sets describing these future scenarios (details in Supplement S2).

## 3. Results

### 3.1 Baseline parameters and sensitivity analysis

The baseline parameter set (Tables 1-3) resulted in a total reduction of the reproduction number R by 12.7%, of which testing and informal tracing (notification of contactees by infected people themselves) contributed 6%, manual tracing (notification by health authorities) another 6.4%, and the contact tracing app only 0.3% (Figure 1a). This was our best estimate of the effectiveness of testing and tracing between December 2020 and March 2021 in the Netherlands. If behaviour is clustered in the population, high-adherence groups are better protected by less circulation in their social network (Figure 1b).

**Figure 1:**
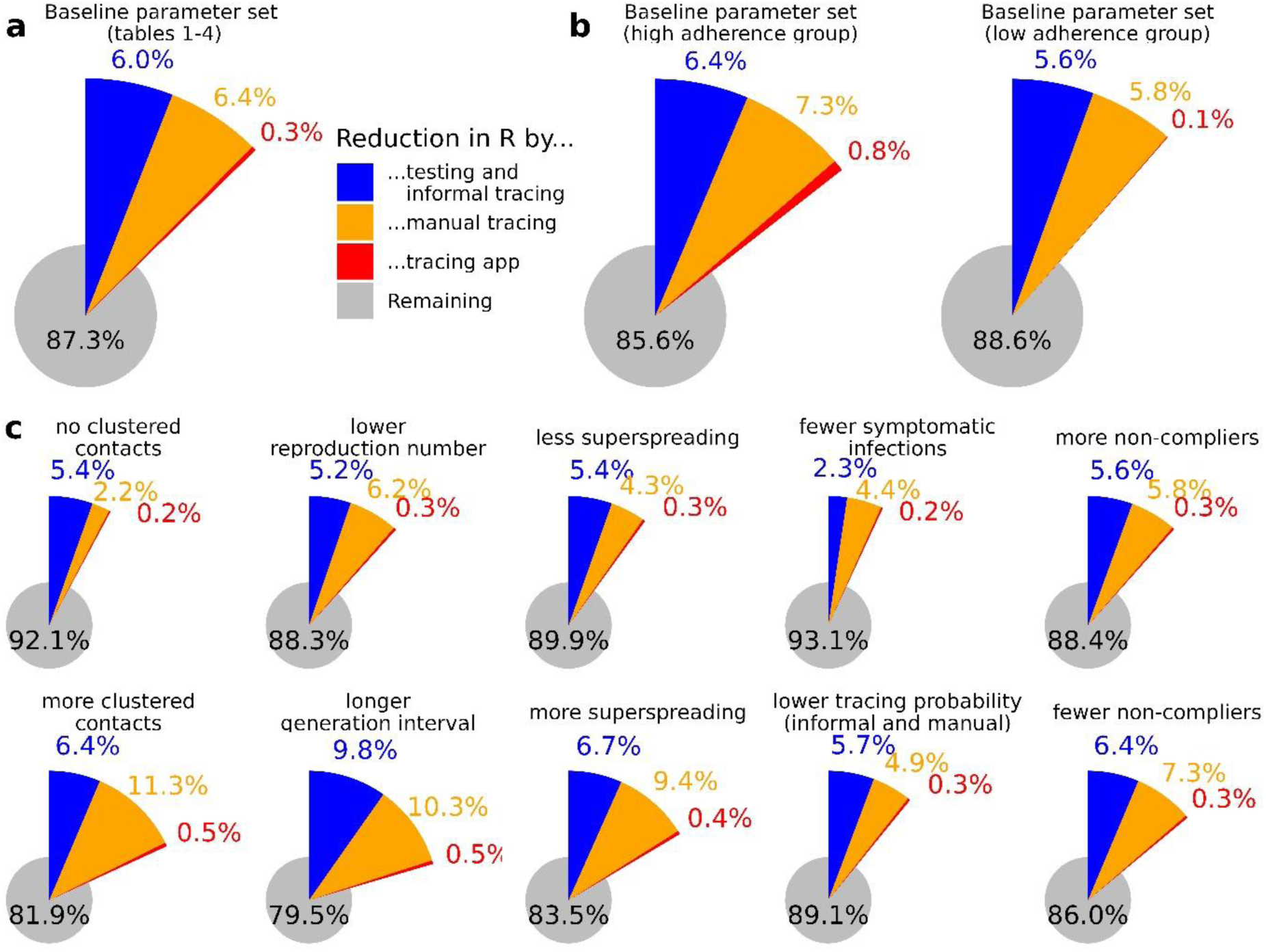
Relative reduction in the reproduction number by testing and tracing, with contributions of the control levels testing and informal tracing, manual tracing, and tracing app; (a) results with the baseline parameter set with parameter values reported in Tables 1-3; (b) results with the baseline parameter set where the population is subdivided into two separate groups with low and high adherence (see sections 2.4 and 2.6) (c) sensitivity analysis with each analysis adjusting exactly one parameter (see section 2.3.5 and Supplement S2 for parameter values)

In Figure 1c the results of the sensitivity analyses of individual parameters with the most uncertainty are presented. The effectiveness of testing and informal tracing did not change very much in most analyses, and became only much less effective with more asymptomatic infections, and more effective with a longer generation interval. Manual tracing effectiveness was very sensitive to changes in the contact network (superspreading and clustering), but also to parameters directly important to the test-and-trace cycle such as the proportion of asymptomatic infection, the probability to trace a contact, and the generation interval which directly affects timely notification of close contacts. Finally, the contribution of app-based tracing remained limited in all sensitivity analyses: 0.2% to 0.5% reduction in *R*.

### 3.2 Scenarios to explore improvements to the app and its use

From the baseline analysis it turned out that the contact tracing app did not contribute much to transmission control. We ran a series of scenario simulations to see if this could change in other conditions. First, we looked at the possibility to change the app such that contact notification can be done by the users themselves, before they are contacted by the health authorities for manual tracing (this became possible later in 2021). This increased the reduction in *R* by app-based tracing from 0.3% to 0.5%, which in relative terms is a lot but not so much in absolute terms (Figure 2a).

**Figure 2:**
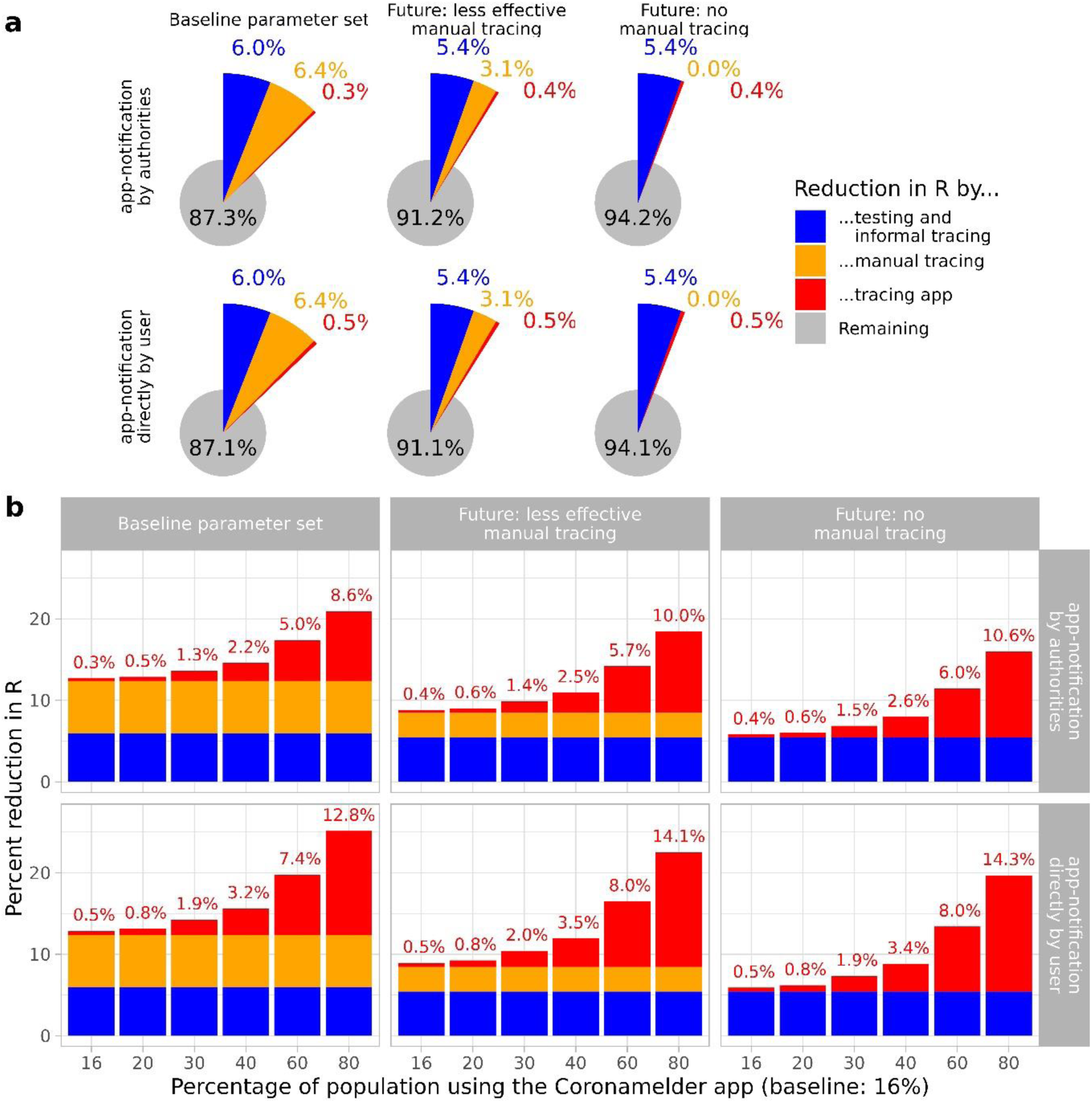
Analysis for future scenarios for the effectiveness of testing, manual tracing, and digital contact tracing in the base setting and a society with (little) restrictions, both with and without formal manual tracing (a) Reduction in the reproduction number, in the top row the app notification is by authorities while in the bottom row the app notification is by the user him/herself (24 vs 8 hours) (b) For the same six parameter sets the effect of a higher percentage app users on the reduction of the reproduction number

Then we looked at how the app would work if lockdown measures were lifted, rendering manual tracing less effective because more contacts are made with strangers, and how it would work when manual tracing would be abandoned at all. It turned out that the reduced contribution of manual tracing was not expected to be compensated by the contact tracing app (Figure 2a).

Finally, we explored how much more effective the contact tracing app would become with more users (Fig 2b), in combination with direct notification by the users and a reduction in manual tracing effectiveness or abandonment of manual tracing. More users made the contact tracing app more effective, but to compensate for the expected lower effectiveness of manual tracing in a future scenario without lockdown, at least 40% of population would need to use the contact tracing app instead of the 16% in the baseline scenario. In that case, the benefit of notification by users themselves also increases, and would contribute 1% of the total reduction of 12.5% (2.5% vs 3.5% - top vs bottom row in Figure 2b, middle column). In the extreme case that 80% of the population would use the contact tracing app, the app could contribute 14.1% of the reduction in *R* (Fig 2b middle column, bottom row).

## 4. Discussion and conclusion

We evaluated the epidemiological effectiveness of the Dutch contact tracing app CoronaMelder in the period from December 2020 to March 2021. We used a simulation model that was informed by testing and tracing data collected in the Netherlands during this study period. We conclude that the contribution of the contact tracing app to control the epidemic was very small, although some protection may have been provided to social groups with more app use. This conclusion was not very sensitive to the major parameter uncertainties. The app could have contributed more if more people would have used it, and if notification of contacts could have been done directly by the user, without involving the health authorities. If formal manual tracing would have become less effective or even abandoned, this would not have been compensated by the app.

The results in our study are at the lower end of the spectrum of effectiveness estimates of contact tracing apps (Jenniskens et al., 2021). It is not straightforward to compare studies, as there are many complexities that play a role (Kretzschmar et al., 2022). Some modelling studies show the effectiveness of the digital contact tracing app in a general setting as a proof of concept, while other studies, like ours, are tailored to a specific country and period (and corresponding policies in place). In particular, the time intervals (Table 3) that were estimated from the data during the evaluation period were relatively long compared to most of the intervals used in literature. It is known that longer time delays in test-and-trace methods can greatly reduce the effectiveness of tracing (Fraser et al., 2004; Klinkenberg et al., 2006; Kretzschmar et al., 2020). In addition, the percentage of app users was low in the Netherlands as compasred to the values used in other studies. Simulations with more users and shorter delays showed the potential benefit of a contact tracing app (Figure 2), which should therefore certainly not be dismissed as a tool in controlling a future epidemic.

Many models for contact tracing have been built, also for evaluation of digital contact tracing during the SARS-CoV-2 pandemic (e.g. (Bradshaw et al., 2021; Cencetti et al., 2021; Kretzschmar et al., 2020; Kucharski et al., 2020; Quilty et al., 2021; Scott et al., 2021)), but our model was unique on three points. First of all, we were able to obtain realistic values for most parameters based on a large variety of data sources of high quality, such as large epidemiological surveillance systems and anonymous questionnaire surveys about behaviour in representative samples of the Dutch population. An important consequence is that our results were a quantitative estimate of the effectiveness of the tracing app in the Netherlands, which could be easily interpreted by policy makers. In fact, the evaluation of the CoronaMelder app (of which the epidemiological evaluation by this model was one part) was followed first by some improvements such as direct notifications by the user, but ultimately by termination of its use (Ministry of Health, 2022).

Second, we made an explicit distinction between formal manual tracing and informal tracing by cases notifying their contacts themselves after receiving a positive test result. Informal tracing was observed in behavioural surveys, and is fast and cheap compared to manual tracing. A proper evaluation of tracing programmes should take this form of tracing into account, because it reduces the additional benefit of manual tracing. On the other hand, manual tracing may reach more contactees and provide the opportunity to better instruct what to do, thus improving the effectiveness of quarantine. In the evaluated context in the Netherlands, manual tracing was estimated to add 6.4% reduction in *R* to the 6% from testing and informal tracing (Figure 1a).

Third, we included the possibility of tracing of contacts that were infected by someone else in the social network, known to be important for the effectiveness of tracing (House and Keeling, 2010; Kretzschmar et al., 2022). This was done by adding contacts to the simulated transmission trees through which tracing could take place. The extent to which these extra contacts are added makes a large difference to the effectiveness of tracing (sensitivity analysis in Figure 1c), which is easily understood as these contacts can bypass the transmission links and lead to quarantine just after or even prior to infection. In our model, we included these contacts so that triangles are created in the original transmission tree, just like triangles are part of clustered networks (Kiss, 2017; Newman, 2003). In clustered networks, the clustering coefficient *c* determines what proportion of triplets form triangles in the network. A topic for further study is how clustering among infected individuals in a transmission tree (as in our model) relates to clustering in an underlying social network.

## Data Availability

All parameter values, code and simulation data used for analysis and visualization are available on Github (https://github.com/rivm-syso/cm-evaluation)

## S1 Model parameters

In this section we provide a detailed description of the sources for each of the parameter values in the study (Tables 1-3)

- Osiris: the national case notification registry containing all confirmed COVID-19 cases (Ward et al., 2005)

- coronIT: all tests carried out by the Municipal Health Services, from all public testing facilities

- HPZone: all contact tracing data

Behavioural data from three large studies were used:

- CGU: questionnaires about beliefs and adherence to measures, ran by the Corona Behavioural Unit of the RIVM (RIVM, 2021)

- LISS: questionnaires about the tracing app, beliefs and adherence to measures, with an existing longitudinal Social Sciences cohort panel (Van der Laan, 2021)

- PanelClix: questionnaire about the tracing app and how it is used, with an existing internet panel (Ebbers, 2021)

Table 1: Parameters of the transmission model Reproduction number *R_ref_*

Value: 1.3

Source: The exact value of R is not essential. A value larger than 1 is necessary to get enough cases in the simulations, to enable calculations. A value close to 1 is representative for the evaluation period under consideration (https://coronadashboard.rijksoverheid.nl/landelijk/reproductiegetal)

Contacts, superspreading coefficient *k* (shape parameter of the negative binomial distribution with mean *R_ref_*)

Value: 0.1

Source: (Kirkegaard and Sneppen, 2021; Susswein, 2020)

Contacts, clustering probability *c*

Value: 0.2

Source: clustering is incorporated into the model because it is known that this influences the effectiveness of infectious disease control through contact tracing (Eames and Keeling, 2003). The implementation of clustering, by adding additional contacts to a transmission tree, is new; we assumed that the clustering probability of our model is similar to the clustering coefficient in population networks. For this, we found estimates of approximately 0.3 for contacts within schools (Kucharski et al., 2018) and 0.1-0.2 from a large scale contact study (Kissler, 2020).

Course of an individual infection, percentage of infections with symptoms

Value: 70%

Source: (Buitrago-Garcia et al., 2020; McDonald et al., 2021)

Course of an individual infection, incubation period

Value: gamma distribution with mean 5 days and standard deviation 2.5 days Source: (Backer et al., 2020; Cheng, 2020)

Course of an individual infection, generation interval

Value: gamma distribution with mean 4 days and standard deviation 2 days

Source: analysis of RIVM Osiris data yields a distribution for the serial interval, from which the distribution of the generation interval can be calculated. The mean coincides between generation interval and serial interval, and for the standard deviation it holds that Var(serial interval) = Var(generation interval) + 2* Var(incubation period)

Table 2: parameters of the behaviour model. Adherence levels of actions in response to triggers Scheduling a test in response to symptoms (own initiative)

Value: 50%

Source: An adherence percentage of 56% with respect to scheduling a test after manual tracing (90%, see below). This 56% is based on the average of people that have gotten tested as estimated from behavioural data ((RIVM, 2021), 10e ronde): “Of the people that have had cold-like symptoms, 46% has gotten themselves tested. Of the people with symptoms that (likely) did not arise from another condition, 57% has gotten themselves tested.”

Scheduling a test in response to informal tracing notification

Value: 68%

Source: We assumed a percentage of 75% with respect to the adherence scheduling a test after manual tracing (90%, see below), which is based on the proportion people that get themselves tested after getting notified by the index him/herself (61.2%) and after notification by the health authorities (83.4%) ((RIVM, 2021) Table S1).

Scheduling a test in response to manual tracing notification

Value: 90%

Source: A percentage of the population (estimated 10%) does not get tested, and as such does not participate in the contact tracing effort. This percentage is an estimate based on the percentage of people that schedules a test after getting notified by the health authorities that they have had close contact with an infectious individual ((RIVM, 2021) Table S1).

Scheduling a test in response to tracing app notification

Value: 81%

Source: We assume a adherence percentage of 90% with respect to scheduling a test after formal manual tracing (90%, see above). We assume the adherence to lie between the percentage of 41% that schedules a test after notification as reported by (Ebbers, 2021) and 95% that has the intention to schedule a test as reported by (Van der Laan, 2021).

Isolation in response to a positive test

Value: 90%

Source: Based on the average percentage between ”staying at home” and “not receiving any visitors” from behavioural data ((RIVM, 2021), 10e ronde): “of the participants that tested positive, 75% report to have stayed at home and 98% reports not having had any visitors”.

Quarantine in anticipation of test outcome, in response to symptoms (own initiative)

Value: 50%

Source: Based on the average between “staying at home” and “not receiving any visitors” based on behavioural data ((RIVM, 2021), 10e ronde): “when people have scheduled a test and have cold-like symptoms that probably didn’t arise from an underlying condition, 34% stayed at home and 70% did not receive visitors” (in round 9 the percentages were 44 and 70%, resp.).

Quarantine in anticipation of test outcome, in response to informal tracing, manual tracing, or tracing app (with symptoms)

Value: 75%

Source: Based on the average percentage between “staying at home” and “not receiving any visitors” from behavioural data ((RIVM, 2021), round 8): “when people are approached by the health authorities in the contact tracing process because they have been in close contact with an infectious individual, 61% stayed at home and 87% did not receive visitors”; “if people received a notification from the health authorities, CoronaMelder app, infectious index him/herself or from school or work, due to close contact, 57% stayed at home and 89% did not receive visitors (round 10)”.

Quarantine in anticipation of test outcome, in response to informal tracing, manual tracing, or tracing app (without symptoms)

Value: 50%

Source: based on (1) Contact tracing app-questionnaire – percentage of people that goes into quarantine when notified by the contact tracing app is 41% (presented as a lower bound (Ebbers 2021) and (2) behavioural data ((RIVM, 2021), round 10): of people that “goes grocery shopping” and “getting fresh air”: “when symptoms (probably) did not arise from an underlying condition, 39% would go out to get fresh air and 32% would go out for grocery shopping. Of the participants with a household member that tested positive, 17% went out to do groceries, 27% went out for fresh air, and 20% went out to walk the dog. Of the ones that were notified by the health authorities, CoronaMelder, infectious index him/herself, or through work or school, 22% went outside for fresh air, 15% went to work, and 15% went grocery shopping.”

Note: the value of 50% adherence to quarantine only applies to those individuals that schedule a test, so the ultimate percentage of individuals that quarantines is lower, as also reported by the behavioural data

Note: without symptoms, after informal notification, the policy was that no test could be scheduled, and therefore the model does not consider quarantine of such individuals.

Table 3: parameters of the logistics model

The time delays in Table 3 are based on the period 1 January until 31 March 2021. In particular, we did not consider the month December 2020 of our evaluation period, due to special events in that month such as holidays. Note that our estimates are optimistic, e.g. we excluded the month December 2020 from the time interval estimation and, due to a lack of data, we assumed that the scheduling of a test follows directly after the notification of a contact from a positive index case.

Below, the rationale behind each time interval parameter in the model is explained.

Delay from symptom onset to scheduling a test (own initiative)

Value: 1.6 days

Source: in Osiris (Ward et al., 2005)the average duration is 2.5 days between symptom onset and positive lab result. The back calculation to the scheduling of the test is through the average duration of 0.5 days between the scheduling of the test and actual taking of the test and 0.4 days between test administration and lab result.

Delay from scheduling a test to test result, with symptoms

Value: 1.3 days

Source: in CoronIT the average time is 0.5 days between scheduling of a test and lab outcome, and 0.4 days between test administration and lab outcome. In addition to those intervals, coronIT yields an average time of 0.4 days between lab outcome and notification of the health authorities, through which a tested individual would get notified.

Delay from scheduling a test to test result, without symptoms

Value: 2.3 days

Source: in CoronIT the average time is 1.5 days between scheduling of a test and test administration, which is 1.0 days longer than the same interval for those with symptoms. In addition, from CoronIT, the time intervals are 0.4 days between test administration and lab result and 0.4 days between lab result and notification of the health authorities.

Delay from positive test result to scheduling a test of a contactee, found by informal tracing notification

Value: 4 hours

Source: no data available. The time interval should be shorter than through tracing app notification (1 day, see below), and is through the social network of the index case, so it should be fast.

Delay from positive test result to scheduling a test of a contactee, found by manual tracing notification

Value: 1.5 days

Source: from HPZone we calculate the time between the notification of the health authorities of the index case and the start date of monitoring a contact. In this calculation we excluded the contacts that were never reached.

Delay from positive test result to scheduling a test of a contactee, found by tracing app notification

Value: 1 day

Source: no data available. The time interval should be shorter than through manual tracing (1.5 days, see above). Because the index still needs to be approached by the health authorities before the tracing app is activated, it cannot be much shorter.

Percentage of infected contactees notified through manual tracing

Value: 40% (31% in figure 3C)

Source: based on the percentage of 40% of cases that is found through manual tracing as reported in Osiris (between 1 January – 31 March 2021). The percentage is equal to the probability of tracing a contact if we assume that the proportion is constant between those infected individuals that do not get tested, those that get tested after formal manual tracing, and those that get tested after symptoms.

Percentage of infected contactees notified through informal tracing

Value: 32%

Source: we assume that the percentage is 80% of the percentage through manual tracing

Percentage of infected contactees notified through tracing app

Value: 75%

- Source: percentage taken from the reported value on the website of the Dutch contact tracing app coronamelder.nl (Note: website is no longer in use. Date at which the url was last accessed: 31 March 2021) See also (Ministry of Health, 2020, 2021a).

Percentage of app users

Value: 16%

Source: the number of app downloads was 4.5 million on 27 January 2021, as percentage of the Dutch population that is 25% (Ministry of Health, 2021b). The estimated percentage of active users is 65% (Ebbers, 2021)

## S2 Parameter sets

**Table S1:** Data from the Corona Behavioural Unit (CGU) (translated from the RIVM website, round 11, 24-28 March: https://www.rivm.nl/gedragsonderzoek/maatregelen-welbevinden/resultaten-11e-ronde-gedragsonderzoek/naleven-gedragsregels)

| Description | % tested total | % tested after close contact | % tested after not-close contact | % stayed at home | % no visitors |
| --- | --- | --- | --- | --- | --- |
| After notification by the index | 61,2<br>(n= 2197) | 84,9<br>(n= 795) | 47,6<br>(n=1380) | 61,5<br>(n= 797) | 89,8<br>(n= 797) |
| After notification by CoronaMelder | 75,7<br>(n= 515) | 94,1<br>(n= 152) | 68,2<br>(n= 258) | 68<br>(n= 153) | 90,8<br>(n= 153) |
| After notification by health authorities/marked as contact by health authorities | 83,4<br>(n= 763) | 91,9<br>(n= 468) | 69,1<br>(n= 285) | 63,9<br>(n= 468) | 90,6<br>(n= 468) |
| After notification through other sources (e.g. school or employer) | 52,4<br>(n= 1089) | 79,8<br>(n= 242) | 43,5<br>(n= 817) | 45,9<br>(n= 242) | 85,1<br>(n= 242) |
| TOTAL (all notifications) | 61,7<br>(n= 3942) | 85,2<br>(n= 1306) | 48,9<br>(n= 2481) | 60<br>(n= 1309) | 89,2<br>(n= 1309) |

**Table S2.** The parameter sets used in the sensitivity analyses of Figure 1 and future scenarios of Figure 2. Parameter values that were different from the baseline set of Tables 1-3 are reported for <u>each parameter set.</u>

| Figure | Description | Parameter value changes compared to Tables 1 - 3 |
| --- | --- | --- |
| 1a | Baseline set | None |
| 1b | High adherence group | Adherence level of scheduling a test in response to<br>- symptoms = 53%<br>- informal notification = 71%<br>- manual notification = 95%<br>- tracing app notification = 86%<br>Percentage app users = 24% |
| 1b | Low adherence group | Adherence level of scheduling a test in response to<br>- symptoms = 48%<br>- informal notification = 64%<br>- manual notification = 85%<br>- tracing app notification = 77%<br>Percentage app users = 8% |
| 1c | No clustered contacts | Clustering probability $c = 0$ |
| 1c | More clustered contacts | Clustering probability $c = 0.3$ |
| 1c | Lower reproduction number | Reproduction number $R_{ref} = 1.05$ |
| 1c | Longer generation interval | Mean generation interval = 5 days |
| 1c | Less superspreading | Superspreading coefficient $k = 0.05$ |
| 1c | More superspreading | Superspreading coefficient $k = 0.5$ |
| 1c | Fewer symptomatic infections | Percentage of infections with symptoms = 30% |
| 1c | Lower tracing probability | Percentage of infected contactees notified through<br>- informal tracing = 25%<br>- manual tracing = 31% (data from CoronIT) |
| 1c | More non-compliers | Adherence level of scheduling a test in response to<br>- symptoms = 48%<br>- informal notification = 64%<br>- manual notification = 85%<br>- tracing app notification = 77% |
| 1c | Fewer non-compliers | Adherence level of scheduling a test in response to<br>- symptoms = 53%<br>- informal notification = 71%<br>- manual notification = 95%<br>- tracing app notification = 86% |
| 2a, b | App notification by authorities | None |
| 2a, b | App notification directly by user | Delay from positive test result to scheduling a test of a contactee, found by tracing app notification = 8 hours |
| 2a, b | Future: less effective manual tracing | Percentage of infected contactees notified through<br>- informal tracing = 16%<br>- manual tracing = 20% |
| 2a, b | Future: no manual tracing | Percentage of infected contactees notified through<br>- informal tracing = 16%<br>- manual tracing = 0% |
| 2b | Percentage of the population using the CoronaMelder app | Percentage of app users = 20%, 30%, 40%, 60%, 80% |

